# BNT162b2 Vaccination efficacy is marginally affected by the SARS-CoV-2 B.1.351 variant in fully vaccinated individuals

**DOI:** 10.1101/2021.07.20.21260833

**Authors:** Orna Mor, Neta S. Zuckerman, Itay Hazan, Ronen Fluss, Nachman Ash, Netanel Ginish, Ella Mendelson, Sharon Alroy-Preis, Laurence Freedman, Amit Huppert

## Abstract

**Background:** Israeli has vaccinated over 80% of its adult population, with two doses of the Pfizer BNT162b2 vaccine. This intervention has been highly successful in curtailing the coronavirus 2 outbreak. One major concern is the ability of the virus to mutate which potentially can cause SARS-CoV-2 to partially escape from the immune system. Here we evaluate the efficacy of the Pfizer vaccine against the B.1.351 variant.

**Methods:** The Ministry of Health, initiated sequencing of selected positive swab samples identified as being of interest. We used logistic regression, with variant type as the dependent variable, vaccination status as the main explanatory variable, controlling for age, sex, subpopulation, place of residence and time of sample, to estimate the odds ratio for a vaccinated case to have the B. 1.351 versus the B.1.1.7 variant, within vaccinated and unvaccinated persons who tested positive.

**Findings:** There were 19 cases of B.1.351 variant (3.2%) among those vaccinated more than 14 days before the positive sample and 88 (3.5%) among the unvaccinated. The estimated odds ratio was 1.29 [95% CI: 0.66-2.50]. From this result, assuming the efficacy against the B.1.1.7 variant to be 95%, the estimated efficacy against the B.1.351 variant was 94% [95% CI: 87-97%].

**Interpretation:** Despite the concerns caused by the B.1.351 variant, the BNT162b2 vaccine seems to provide substantial immunity against both that variant and the B.1.1.7. Our results suggest that from 14 days following the second vaccine dose the efficacy of BNT162b2 vaccine is at most marginally affected by the B.1.351 variant.

**Funding:** No funding

## Introduction

The impressive success of mass vaccination on curtailing the coronavirus 2 (SARS-CoV-2) by halting transmission has paved the road for returning to “pre-pandemic life”. However, several open questions challenge the triumph of controlling/ending the pandemic by vaccination. One major concern is the ability of the virus to mutate and evolve; this potentially can cause SARS-CoV-2 to partially escape from the immune system, which will reduce the effectiveness of the vaccine in preventing disease and viral transmission. Determining vaccine efficacy against variants of concern (VOC)^1^ is vital for planning and modifying vaccination strategies.

On December 19^th^ 2020, Israel launched a massive COVID vaccination campaign based on the Pfizer BNT162b2 vaccine, and by end of May 2021, had administered over 10,500,000 doses, to approximately 5,400,000 individuals, more than 80% of the population over 16y, receiving two doses. Both in clinical trials and in real world studies, the BNT162b2 vaccine has proven to be highly effective in both averting infections and preventing severe disease and death. ^2-5^

The Israeli vaccination campaign took place during the third and largest wave of the pandemic (see Figure 1). During this third wave, the B.1.1.7 variant became the dominant strain in Israel, reaching over 95% dominance. ^6^ Since the detection of the B.1.1.7 variant in November 2020 in the United Kingdom, it has spread rapidly and become the dominant strain in many countries. There is also evidence that it causes higher rates of morbidity and mortality. ^7^ Nevertheless, the BNT162b2 vaccine, which was developed based on the original Wuhan strain sequence, has been found very effective against the B.1.1.7 variant, both in blocking transmission and reducing morbidity and mortality following infection. ^2–5^ The B.1.351 strain, which was first documented in South Africa, is also considered a VOC mainly because *in vitro* experiments have demonstrated its ability to overcome previous immunity to SARS-CoV2. Specifically, experimental work demonstrated significant decrease in neutralization capacity of B.1.351. ^8^ However other research found that neutralizing antibodies remained sufficiently high against the B.1.351 variant. ^9^ Humoral protection measured by antibody responses and neutralization studies do not assess the role of cellular immunity mediated by T-cell responses. A recent study has shown that the cellular protection established following previous infection or vaccination remains high against both the B.1.1.7 and the B.1.351 variants.^10^ On the other hand, two real world studies have raised concern that the BNT162b2 vaccine has reduced efficacy against the B.1.351 variant. A study from Qatar has shown that the efficacy of BNT162b2 against the B.1.351 variant was ∼75% compared to ∼90% against the B.1.1.7 variant. ^11^ A second study from Israel has estimated that the odds ratio (OR), in a matched study of SARS-Cov-2 cases occurring in unvaccinated persons versus persons who had received their second dose of vaccine at least one week previous to sample collection, was 1/8, implying considerably lower efficacy of the vaccine against the B.1.351 variant. ^6^ Thus, our goal was to further quantify the risk of the B.1.135 variant causing a significant breakthrough in a real world environment.

**Figure 1:**
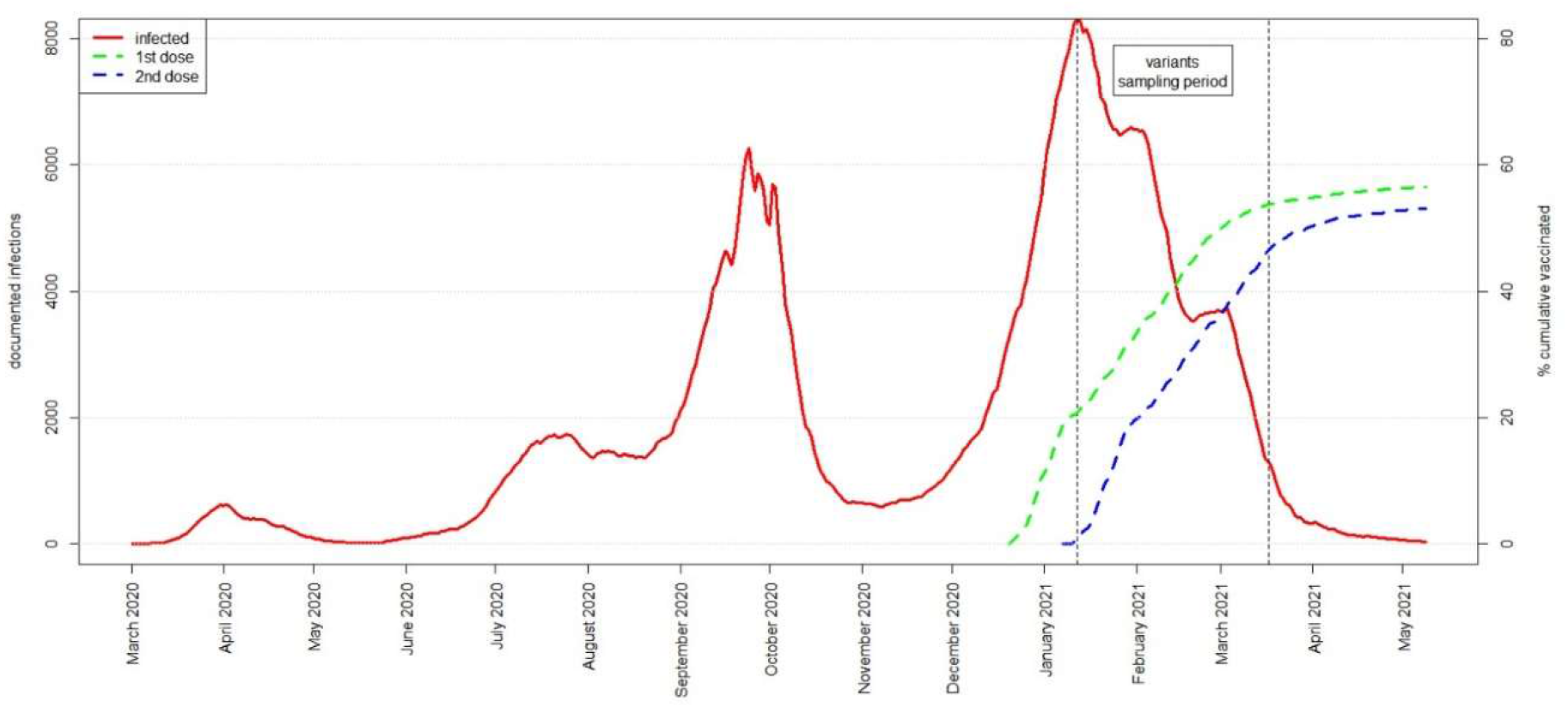
Number of documented COVID19 infections in Israel from March 2020 until May 2021 indicated by the red line. The Green and blue dashed lines represent the percent of the population vaccinated with the first and second doses respectively.

## Methods

### Collection of samples

With the start of the vaccination campaign in Israel, the Central Virology Laboratory (CVL) of the Ministry of Health, initiated collection and sequencing of selected swab samples that had tested positive on polymerase chain reaction (PCR). Samples were selected for sequencing *i*) to monitor the circulating and imported variants in Israel *ii*) to characterize viral variants among cases occurring after vaccination and matching cases in unvaccinated persons *iii*) to monitor local outbreaks, severe clinical cases and transmission among specific population groups, and *iv)* to follow-up on those coming into contact with persons found to have the B.1.351 variant. Samples identified as being of interest were retrieved from the 48 laboratories that perform SARS-CoV-2 PCR tests in Israel, and sent to CVL, where they were assessed by whole genome sequencing.

### Laboratory methods

RNA extracted from SARS-CoV-2 positive samples was sequenced with COVID-Seq library preparation on NovaSeq (Sp 300 cycles, Illumina, CA, USA). Resulting fastq files were processed, including quality filtering, mapping to the reference genome (NC_045512.2), construction of consensus fasta sequences, alignment to the reference genome and mutation analyses via a custom python-based pipeline. Specific variants were determined based on identification of relevant mutations for each variant (B.1.1.7 [doi: 10.1016/j.jinf.2020.12.024], B.1.351 [doi: https://doi.org/10.1101/2020.12.21.20248640]) in the genome sequence and identification of the relevant lineage by Pangolin [doi:10.1038/s41564-020-0770-5]. Additional information provided in the supplementary.

### Creation of the database

The information on all cases sent for sequencing was entered in a database, containing: socio-demographical information (e.g. age, city/town/village of residence, subpopulation - Arab, Ultra-Orthodox Jewish, other), date of collection of first positive sample, date of recovery, vaccination dates, symptoms and hospitalizations, and the infecting variant as determined by whole genome sequencing. All information was retrieved from the Israeli Ministry of Health’s databases.

### Statistical methods

We restricted our analyses to vaccinated and unvaccinated cases that were positive either for the variant B.1.1.7 or the variant B.1.351 using whole genome sequencing. Vaccinated cases were defined as those where the first positive sample was taken at least 14 days after the second dose. Those with unknown dates of vaccination, or who received only one vaccine dose, or for whom the sample was taken between the first dose and second dose, were excluded.

Those for whom the sample was taken less than 14 days after their second dose were excluded from the main analysis, but were included in a secondary analysis. Individuals who had acquired the infection outside Israel, those aged less than 16y (not eligible for vaccination) and individuals without information regarding their place of residence, were all excluded.

The main principle of the statistical analysis was to estimate the odds ratio, OR, for a vaccinated case to have the B.1.351 variant, within vaccinated and unvaccinated persons who have tested positive. One can show (see Supplementary Information) that:

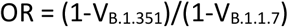

where V _B.1.351_ is the vaccine efficacy against the B.1.351 variant, and V_B.1.1.7_ is the vaccine efficacy against the B.1.1.7 variant. Thus, assuming that V_B.1.1.7_ is estimated well from observational studies of vaccine efficacy conducted in Israel (e.g., 95%), we estimated the vaccine efficacy against the B.1.351 variant from:

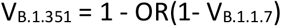

From this equation, when OR equals 1, then V_B.1.351_ = V_B.1.1.7_. Thus, a test of the hypothesis that OR = 1, also tests whether the vaccine is equally effective against each variant.

We estimated the odds ratio by logistic regression^12^, with variant type as the dependent variable and vaccination status as the main explanatory variable. The following variables were entered as potential confounding covariates: city/town/village of residence (as a random effect), date of taking the swab sample (in five categories: 20-31 Dec 2020; 1-31 Jan 2021; 1-14 Feb 2021; 15-28 Feb 2021; 1-31 Mar 2021), subpopulation (Arab, Jewish ultra-orthodox, other), and age group (16-44, 45-64, 65-79, ≥80y). The analysis was implemented using the glmer procedure in the lme4 package of the R software.^13^ This approach also provided a p-value for the test that OR=1 and a 95% confidence interval for the OR.

A secondary analysis was conducted to examine the influence of time of infection following full vaccination, using the same methods as described above and comparing unvaccinated cases versus cases where the sample from the vaccinated case was taken within the first 14 days after the second dose of vaccination.

Alternative analyses were conducted using matching to control for confounding variables, and are reported in the Supplementary Information. Unlike the main analyses presented here, such matching entails exclusion of approximately 60% of the cases. We regard their results as providing information supportive to the main analyses.

## Results

The database contained the sequencing results of 11624 samples obtained from distinct individuals. After the exclusions described in the Methods section, 596 vaccinated and 2515 unvaccinated cases were left eligible for analysis (Figure 2). Characteristics of these vaccinated and unvaccinated individuals are shown in Table 1. The vaccinated groups were on average older and had a smaller proportion of ultra-orthodox Jews, emphasizing the need to control for these potential confounders.

**Figure 2:**
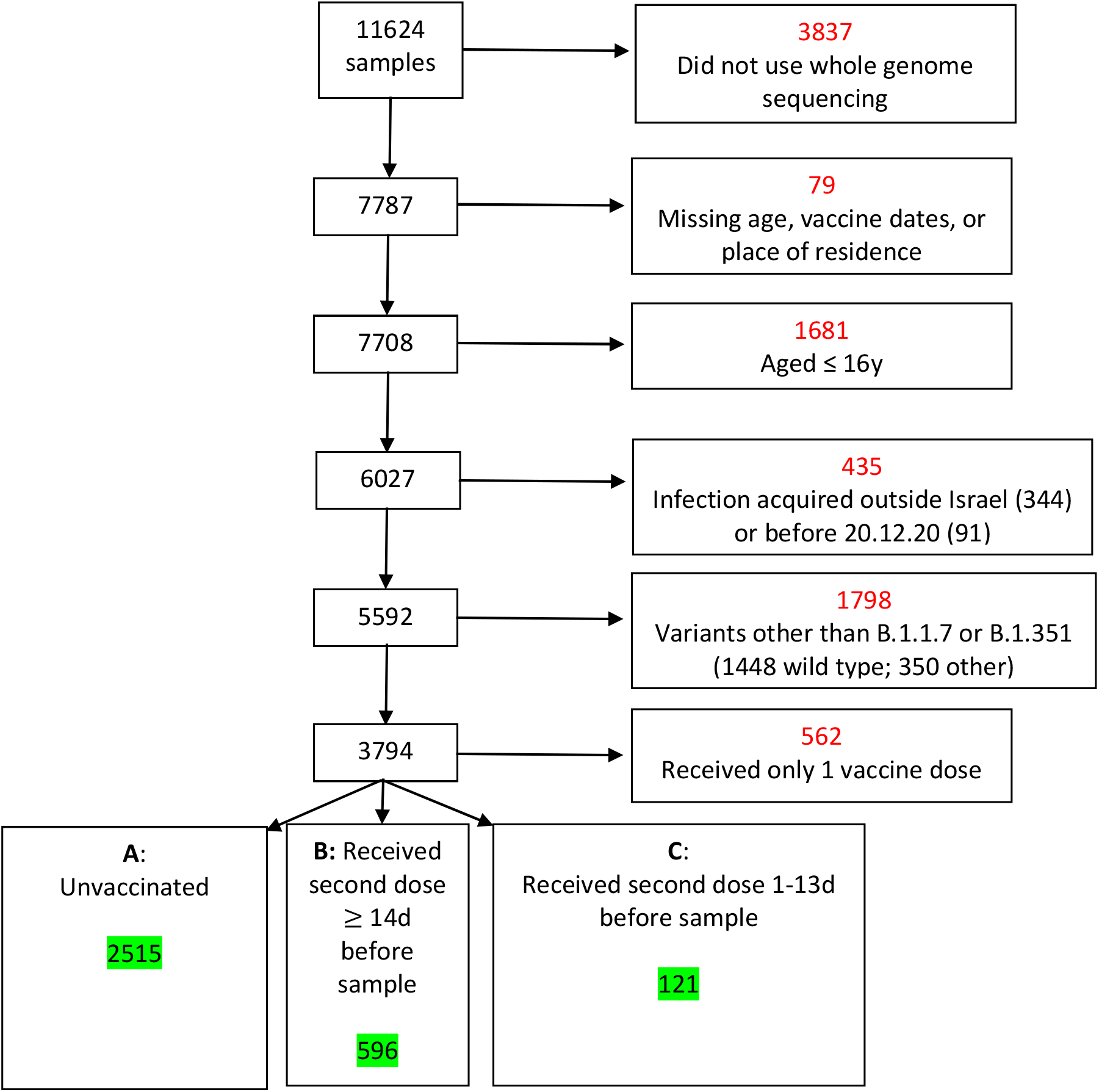
Flow chart showing number of exclusions (in red) from the analysis, with the reasons for exclusion. Numbers included in the analyses are shown in green.

**Table 1:**
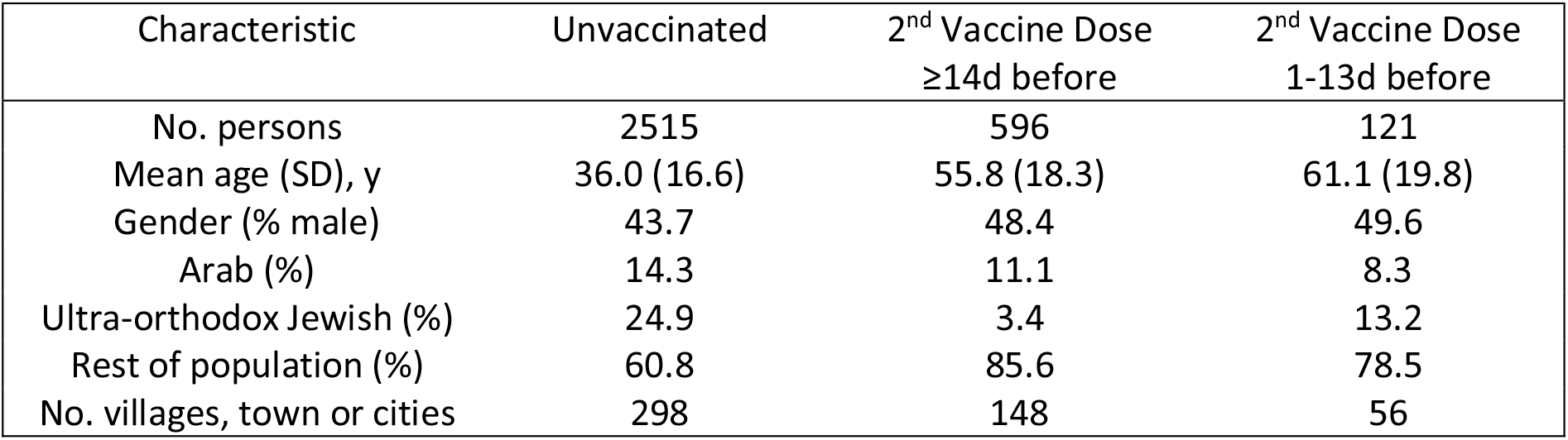
Characteristics of those included in the analysis

| Characteristic | Unvaccinated | 2<sup>nd</sup> Vaccine Dose<br>≥14d before | 2<sup>nd</sup> Vaccine Dose<br>1-13d before |
| --- | --- | --- | --- |
| No. persons | 2515 | 596 | 121 |
| Mean age (SD), y | 36.0 (16.6) | 55.8 (18.3) | 61.1 (19.8) |
| Gender (% male) | 43.7 | 48.4 | 49.6 |
| Arab (%) | 14.3 | 11.1 | 8.3 |
| Ultra-orthodox Jewish (%) | 24.9 | 3.4 | 13.2 |
| Rest of population (%) | 60.8 | 85.6 | 78.5 |
| No. villages, town or cities | 298 | 148 | 56 |

The distribution of variants (B.1.1.7 and B.1.351) by vaccination status is shown in Table 2. There were 19 cases of B.1.351 variant (3.2%) among those vaccinated more than 14d before the positive sample and 88 (3.5%) among the unvaccinated. The estimated OR (Table 3) was 1.29, with a P-value of 0.46 and 95% confidence limits ranging from 0.66 to 2.50. Note that the estimated OR was larger than one even though the crude proportion of B.1.351 was slightly lower (3.2% v 3.5%) among the vaccinated cases. This was due to the regression adjustment for the confounders age and subpopulation – younger age and ultra-orthodox Jews both had a negative association with the B.1.351 variant. Using the estimated OR, assuming that the vaccine efficacy against the B.1.1.7 variant is 95%^2-5^, the estimated efficacy against the B.1.351 variant is estimated to be 93% with 95% confidence limits between 87% and 97%. Results from the supportive analysis when matching was employed gave an estimated OR of 0.90 (95%CI: 0.40-2.01) (see Supplementary Tables A1-A4 for details).

**Table 2:** Distribution of variants among vaccinated and unvaccinated cases

| Variant | Unvaccinated | 2<sup>nd</sup> Vaccine Dose<br>≥14d before | 2<sup>nd</sup> Vaccine Dose<br>1-13d before |
| --- | --- | --- | --- |
| B.1.351 | 88 (3.5%) | 19 (3.2%) | 14 (11.6%) |
| B.1.1.7 | 2427 | 577 | 107 |
| Total | 2515 | 596 | 121 |

**Table 3:** Estimated odds ratio and vaccine efficacy against B.1.351 variant

| Estimated Odds Ratio | 95% CI | P-value | Vaccine Efficacy against B.1.351 variant (95% CI)<sup>1</sup> |  |
| --- | --- | --- | --- | --- |
| | | | $V_{B.1.1.7} = 90\%$ | $V_{B.1.1.7} = 95\%$ |
| 1.29 | 0.66-2.50 | 0.46 | 87% (75-93) | 94% (87-97) |

<sup>1</sup> Calculated from $1 - (\text{Odds Ratio} \times (1 - V_{B.1.1.7}))$

The same methods as above were applied to persons who received their second dose between 1 and 13 days before collection of the sample. In the 121 cases, there were 14 (11.1%) B.1.351 variants; the estimated OR was 2.62 (P=0.015, 95%CI = 1.20-5.70). A similar estimate was obtained from the supportive analysis using matching, but with a wider confidence interval (estimated OR = 2.12, 95%CI = 0.49-9.05).

## Discussion

The great success of the Israeli vaccination campaign against SARS-CoV-2 can be appreciated by the fact that, as of June 1^st^, Israel lifted all COVID emergency restrictions except the requirement to wear masks indoors and regulations governing international travel.

Variants can affect a vaccine’s impact on the transmission of an infection by two factors. First, VOC may have a higher rate of transmission that will require a higher vaccination coverage or a greater vaccine efficacy to curtail the spread of infection.^14^ In this regard, the B.1.1.7 variant is about 50% more infectious than the original Wuhan strain^7^, leading to the subsequent global dominance of B.1.1.7. This higher transmission rate requires higher vaccine coverage to allow relaxation of social distancing and other non-pharmaceutical interventions while still curtailing new outbreaks. The second factor is vaccine breakthrough by VOC. Initially, evidence regarding the evolutionary dynamics governed by the relatively slow mutation rate bolstered the hope that VOC would not endanger the effectiveness of vaccination. However, concerns over reduced efficacy of the Pfizer BNT162b2 vaccine arose after laboratory experiments estimated a large drop in sensitivity of the B.1.1.7 variant to sera collected from vaccinated individuals and an even larger reduction in sensitivity to convalescent sera.^8^ The B.1.351 variant has caused further anxiety regarding the potential vaccine breakthrough due to the co-occurrence of supplementary mutations in the receptor-binding domain, which were proven to have a significantly increased resistance to vaccine-induced and convalescent sera.^15^ Two real-world studies which examined the protection of the BNT162b2 vaccine against B.1.1.7 and B.1.351 found a substantial reduction in vaccine effectiveness against B.1.351 compared to B.1.1.7. ^6,11^

Results from a previous study in Israel based on a matched pairs case-case design pointed to a significant vaccine breakthrough of the B.1.351 compared to the B.1.1.7. The study had a modest sample size and period-length, and found eight B.1.351 breakthroughs compared to a single case of a B.1.17 breakthrough in the subgroup of nine discordant matched pairs (i.e., those for which the vaccinated and unvaccinated cases differed in the variant-type) of vaccinated persons receiving two vaccine doses and unvaccinated persons;^6^ all these cases were PCR confirmed in a short time window, on days 7-13 after receiving the second dose. No positive individuals infected by the B.1.351 variant were found in vaccinated persons more than two weeks after the second dose even though about half of the vaccinated cases were from that later time frame. The investigators speculated that “this cohort may have been infected before the immunity from the boost was fully established, and it is thus possible that enhanced immunity from the boost, which develops over time, may more effectively prevent infection with the B.1.351 variant.”

In our study, a much larger number of vaccinated and unvaccinated cases was included. In addition, our main analysis focused on vaccinated cases infected with B.1.351 or B.1.1.7 occurring more than two weeks after their second dose and did not find a statistically significant reduction in protection of the vaccine. However, a sub-analysis of our study, which examined vaccinated cases occurring only 1-13 days after the second dose found an increased proportion of B.1.351 compared to unvaccinated cases with an estimated OR of 2.62. It is important to mention that, although both studies were conducted in Israel, there is no overlap of data between the two studies. Integrating the information and findings from both studies, supports the hypothesis that a higher level of immunity is required for protection against the B.1.351 variant, thus inducing different levels of efficacy against the B.1.351 and B.1.1.7 variants during the first two weeks after the boost from the second dose.^16^ The fact that the B.1.351 variant, that was first diagnosed in Israel January 2021, concomitantly with the start of the vaccination program, has not caused significant community transmission in Israel is further encouraging evidence supporting the ability of two doses of the BNT162b2 vaccine combined with high coverage to halt transmission.

A second study used a test-negative design to examine the efficacy of the BNT162b2 vaccine in Qatar,^11^ and found that the vaccine efficacy was 89.5% (95% CI:[85.9-92.3]) against the B.1.1.7 variant but only 75% (95% CI:[70.5-78.9]) against the B.1.351 variant, at least 14 days after the second dose. In contrast, several large studies from Israel have shown that the efficacy of the BNT162b2 vaccine is close to 95% against infection during a time when the B.1.1.7 variant was predominant. ^3–5^ The difference in vaccine efficacies to the B.1.1.7 variant between the Qatar and Israeli studies could be due to the use of different methodologies to estimate vaccine efficacy, or different social, cultural, racial or environmental conditions; the latter potentially can affect the transmission dynamics and thus alter vaccine efficacy. The above differences could also explain the difference in vaccine breakthrough of the B.1.351 variant between the two countries. The current reduction in transmission seen in Qatar after the vaccination campaign gives further support to the ability of the BNT162b2 vaccine to control the spread of the B.1.351 variant.

Limitations of our study include the relatively low number of B.1.351 cases, owing to its low prevalence in Israel, and also the fact that the sequencing was not done on a sample selected randomly from the total population of SARS-Cov2 positive cases in Israel. In particular, samples were selected for sequencing based on a variety of concerns, but partly to check on the variants among cases occurring after vaccination and partly to follow-up on those coming into contact with persons found to have the B.1.351 variant. Thus, the database has over-representation of vaccinated cases, and also over-representation of the B.1.351 variant among cases in Israel. In order to test the robustness of our results, we used two different statistical methods, unconditional logistic regression and conditional logistic regression with matching to estimate the OR. Both analyses revealed no statistically significant difference, and indicated that any reduction in the vaccine efficacy against B.1.351 relative to B.1.1.7 is at the most marginal with an OR not exceeding 2.5. In addition, we have shown in computer simulations that this type of selection should not introduce bias into the estimation of the odds ratio of a vaccinated case having the B.1.351 variant (see Supplemental Information for details). The criterion for validly using such a database for estimating the OR is that the selection of vaccinated cases or B.1.351 cases should be independent of each other; when selecting a vaccinated case this should be without regard to whether the case was B.1.17 or B.1.351, and when selecting a case suspected to be B.1.351 this should be without regard to whether the case was vaccinated or unvaccinated. As far as we can ascertain such conditions applied to the selection of cases for sequencing.

Despite the concerns caused by the B.1.351 variant, the BNT162b2 vaccine seems to provide substantial immunity against both that variant and the B.1.1.7. Our results suggest that from 14 days following the second vaccine dose the efficacy of BNT162b2 vaccine is at most marginally affected by the B.1.351 variant. The next generation of vaccines will include modifications to better deal with different VOC; nevertheless, the current vaccines may still provide substantial immunity against both current and future VOC.

### Helsinki approval

This study was conducted according to the guidelines of the Declaration of Helsinki, and approved by the Institutional Review Board of the Sheba Medical Center (7045-20-SMC). Patient consent was waived because this study used remains of clinical samples and the analysis used anonymous clinical data.

## Data Availability

Data is the property of the Israeli Ministry of Health and is not available to the public at this stage.

## Supplementary Information

### Supplementary Laboratory Methods

RNA from samples tested positive for SARS-CoV-2 via real-time PCR was extracted from 200 μL respiratory samples with the MagNA PURE 96 (Roche, Mannheim, Germany) according to the manufacturer instructions. Whole genome SARS-CoV-2 sequencing libraries were prepared with COVID-Seq (Illumina, CA, USA) and sequenced on NovaSeq using Sp kit with 300 cycles (Illumina, CA, USA). A custom python-based pipeline used to process the resulting fastq files included quality assessment with FastQC (www.bioinformatics.babraham.ac.uk/projects/fastqc/) and MultiQC [doi: 10.1093/bioinformatics/btw354], filtering low-quality sequences with trimmomatic [doi: 10.1093/bioinformatics/btu170], mapping to SARS-CoV-2 reference genome (NC_045512.2) with Burrows-Wheeler aligner (BWA) [doi: 10.1093/bioinformatics/btp324], filtering unmapped reads, sorting, indexing and determining sample coverages using SAMtools suite [doi: 10.1093/bioinformatics/btp352], alignment to the reference genome with MAFFT [doi: 10.1093/nar/gkf436] and identification of mutations reported to be associated with specific variants, e.g. B.1.1.7 [doi: 10.1016/j.jinf.2020.12.024] and B.1.351 [doi: https://doi.org/10.1101/2020.12.21.20248640]. Validation of lineages was done with Pangolin lineages [doi:10.1038/s41564-020-0770-5].

For supplementary statistical analysis:

### Matching analysis

Each matched set comprised a single vaccinated case matched to one or more (up to 10) unvaccinated cases. Matching on all of the following variables was required: city/town/village of residence, date of taking the swab sample (± 7d), and subpopulation (Arab, Jewish ultra-orthodox, other). We estimated the odds ratio by conditional logistic regression, ^17^ with variant type as the dependent variable and vaccination status as the explanatory variable. The analysis was implemented using the clogit procedure in the survival package of the R language. ^18^ Age group (16-44, 45-64, 65-79, ≥80y) was included instead as a covariate in the regression model. After the exclusions described in the Methods section, 596 vaccinated and 2524 unvaccinated cases were left eligible for analysis. Of these 596 vaccinated cases, matched unvaccinated cases could be found for 422 (71%). These 422 vaccinated cases were matched with 861 unvaccinated cases. The distribution of the number of matched unvaccinated cases per matched set is shown in Table A1, and characteristics of the vaccinated and unvaccinated individuals in these matched sets are shown in Table A2.

Matching on the same variables as the main analysis, but also including age, yielded a total of 290 matched sets, with 290 vaccinated and 563 matched unvaccinated cases. Table A1 shows the distribution of the number of unvaccinated case matches per vaccinated case.

**Table A1:** Number of matched unvaccinated cases to each vaccinated case achieved

| Number of unvaccinated matches per set | Number of matched sets |
| --- | --- |
| 1 | 209 |
| 2 | 121 |
| 3 | 45 |
| 4 | 14 |
| 5 | 11 |
| 6 | 7 |
| 7 | 8 |
| 8 | 1 |
| 9 | 2 |
| 10 | 4 |
| Total | 422 |

**Table A2:** Characteristics of those included in the matched analysis

| Characteristic | Unvaccinated | Vaccinated |
| --- | --- | --- |
| No. persons | 861 | 422 |
| Mean age (SD), y | 37.9 (17.5) | 56.6. (18.4) |
| Gender (% male) | 43.1 | 46.1 |
| Arab (%) | 13.8 | 10.0 |
| Ultra-orthodox Jewish (%) | 7.5 | 4.3 |
| Rest of population (%) | 78.6 | 85.8 |
| No. villages, town or cities | 85 | 85 |

**Table A3:** Distribution of variants among vaccinated and unvaccinated cases: matching that did not include age

| Variant | Vaccinated | Unvaccinated |
| --- | --- | --- |
| B.1.351 | 16 (3.8%) | 25 (2.9%) |
| B.1.1.7 | 406 | 836 |
| Total | 422 | 861 |

**Table A4:** Estimated odds ratio and vaccine efficacy against SA variant: matching did not include age but age-group was adjusted for in the regression model

| Estimated Odds Ratio | 95% CI | P-value | Vaccine Efficacy against B.1.351 variant (95% CI)<sup>1</sup> |  |
| --- | --- | --- | --- | --- |
| | | | $V_{B\ 1.1.7}=90\%$ | $V_{B\ 1.1.7}=95\%$ |
| 0.90 | 0.4-2.0 | 0.79 | 91% (80-96) | 95% (90-98) |

<sup>1</sup> Calculated from $1 - (\text{Odds Ratio} \times (1 - V_{B\ 1.1.7}))$

### Computer simulations to check on effect of selection on the estimate of the odds ratio

The simulation was written in the R language, and the code is given below.

The computer generates a very large study based on sampling preferentially more vaccinated cases and more B.1.351 cases than are representatively found in the population of cases.

However, the preferential selection of vaccinated cases is without reference to whether they have the B.1.351 or the B.1.1.7 variant, and the preferential selection of B.1.351 cases is without reference to whether they are vaccinated or unvaccinated. The selected study data are then used to estimate the odds ratio and this estimate is then compared to the “true odds ratio” that was used to generate the data.

In the following R code notation is as follows:

p0 = probability of infection by B.1.1.7

p1 = probability of infection by B.1.351

v0 = vaccine efficacy against B.1.1.7

v1 = vaccine efficacy against B.1.351

pvac = proportion vaccinated

svac0 = proportion of cases of unvaccinated cases selected for sequencing

svac1 = proportion of cases of vaccinated cases selected for sequencing

svar0 = proportion of cases of B.1.1.7 cases selected for sequencing

svar1 = proportion of cases of B.1.351 cases selected for sequencing

~~~
vari.sim3
function(nsim,n,p0,p1,v0,v1,pvac,svac0,svac1,svar0,svar1){
  n00<-rep(0,nsim)
  n01<-rep(0,nsim)
  n10<-rep(0,nsim)
  n11<-rep(0,nsim)
  nsel00<-rep(0,nsim)
  nsel01<-rep(0,nsim)
  nsel10<-rep(0,nsim)
  nsel11<-rep(0,nsim)
  for(i in 1:nsim) {
vac<-rbinom(n,1,pvac)
pdis0<-p0*(1-vac*v0)
pdis1<-p1*(1-vac*v1)
dis0<-rbinom(n,1,pdis0)
dis1<-rbinom(n,1,pdis1)
pselvac <- vac*svac1 + (1-vac)*svac0
pselvar <- dis0*svar0*(1-dis1) + dis1*svar1*(1-dis0) psel <- pselvar*pselvac
sel <- rbinom(n,1,psel)
n00[i]<-sum(dis0==1 & vac==0)
n01[i]<-sum(dis0==1 & vac==1)
n10[i]<-sum(dis1==1 & vac==0)
n11[i]<-sum(dis1==1 & vac==1)
nsel00[i]<-sum(sel==1 & dis0==1 & vac==0)
nsel01[i]<-sum(sel==1 & dis0==1 & vac==1)
nsel10[i]<-sum(sel==1 & dis1==1 & vac==0
nsel11[i]<-sum(sel==1 & dis1==1 & vac==1)
}
c(sum(n00),sum(n01),sum(n10),sum(n11),sum(n11)*sum(n00)/(sum(n10)*sum(n
01)),sum(nsel00),sum(nsel01),sum(nsel10),sum(nsel11),sum(nsel11)*sum(ns
el00)/(sum(nsel10)*sum(nsel01)))
}
~~~

In the above program, the term `sum(n11)*sum(n00)/(sum(n10)*sum(n01))` is the odds ratio in the population and estimates the ratio (1-v1)/(1-v0).

The term `sum(nsel11)*sum(nsel00)/(sum(nsel10)*sum(nsel01))` is the odds ratio in the cases selected for screening.

The following table gives the results of running this program for different vaccine efficacies, and selection proportions.

**Table A4:** Results of computer simulation comparing the estimated odds ratio in the full population of samples and the population selected for sequencing

| v0 | v1 | True OR | Population<br>estimated<br>OR | svac0 | svac1 | svar0 | svar1 | Selected<br>sample<br>estimated<br>OR |
| --- | --- | --- | --- | --- | --- | --- | --- | --- |
| 0.9 | 0.9 | 1.0 | 1.01 | 0.6 | 1.0 | 0.2 | 1.0 | 1.03 |
|  |  |  | 1.01 | 0.5 | 1.0 | 0.1 | 1.0 | 1.01 |
|  |  |  | 1.01 | 0.2 | 1.0 | 0.05 | 1.0 | 1.06 |
| 0.9 | 0.8 | 2.0 | 2.03 | 0.6 | 1.0 | 0.2 | 1.0 | 2.03 |
|  |  |  | 1.98 | 0.5 | 1.0 | 0.1 | 1.0 | 2.03 |
|  |  |  | 2.03 | 0.2 | 1.0 | 0.05 | 1.0 | 2.07 |
| 0.9 | 0.6 | 4.0 | 3.98 | 0.6 | 1.0 | 0.2 | 1.0 | 3.96 |
|  |  |  | 4.06 | 0.5 | 1.0 | 0.1 | 1.0 | 4.08 |
|  |  |  | 3.97 | 0.2 | 1.0 | 0.05 | 1.0 | 3.98 |

Comparing the final column of the table with the true odds ratio (3^rd^ column) or the estimated OR in the population, one can see that the estimated odds ratio in the selected sample appears to have little or no bias.

### Demonstration of the relationship between vaccine efficacy and the odds ratio in a matched case-case design

The following theory shows that estimating the odds ratio of having the B.1.351 variant among the vaccinated to unvaccinated cases will allow one to estimate the efficacy of the vaccine against the B.1.351 variant relative to the efficacy of the vaccine against the B.1.1.7.

Consider a matched pair.

Assume the risks of catching each variant conditional on the matching covariates and no vaccination are as follows:

P(B.1.1.7 variant | covariates, no vaccine) = π_*B.1.1.7*_

P(B.1.351 variant | covariates, no vaccine) = π_*B.1.351*_

Among such persons who get either the B.1.1.7 or B.1.351 variant, the proportion with the

B.1.351 variant will be

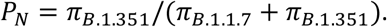

Let the vaccine efficacy against the B.1.1.7 variant be *V*_B.1.1.7_, and against the B.1.351 variant

*V*_B.1.351_. Among the same type of persons who are vaccinated:

P(B.1.1.7 variant | covariates, vaccine) = π_*B.1.1.7*_(1 − *V*_*B.1.1.7*_)

P(B.1.351 variant | covariates, vaccine) = π_*B.1.351*_(1 − *V*_*B.1.351*_)

Among such persons who get COVID-19, the proportion with the B.1.351 variant will be

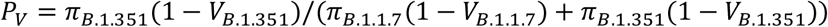

So for this type of person, the odds ratio of having the B.1.351 variant for vaccinated versus unvaccinated is:

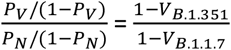

## Notes

### Competing Interest Statement

The authors have declared no competing interest.

